# Anxiety due to Long COVID is partially driven by activation of the tryptophan catabolite (TRYCAT) pathway

**DOI:** 10.1101/2023.04.24.23288115

**Authors:** Hussein Kadhem Al-Hakeim, Anwar Khairi Abed, Abbas F. Almulla, Shatha Rouf Moustafa, Michael Maes

## Abstract

This study examines whether activation of the tryptophan catabolite (TRYCAT) pathway is associated with anxiety symptoms due to Long COVID. We selected 90 participants, 60 Long COVID patients and 30 individuals without any symptoms following acute CIVID-19 infection. Using cluster analysis and the Hamilton Anxiety Rating scale (HAMA) score, the pure HAMA anxiety score, serum tryptophan (TRP) and kynurenine (KYN), the KYN/TRP ratio (all measured during Long COVID), and oxygen saturation (SpO2) (measured during the acute phase of COVID-19), we were able to classify Long COVID patients into two distinct clusters with an adequate silhouette cohesion and separation index (0.58): cluster 1 (n=61) and cluster 2 (n=29). Cluster 2 patients had lower SpO2 and TRP levels, as well as higher KYN, KYN/TRP ratio, and HAMA scores than cluster 1. Regression analysis revealed that the KYN/TRP ratio explained 14.4% of the variance in the HAMA score (F=14.81, df=1/88, p=0.001). In addition, regression analysis revealed that SpO2 partially explained the variance in serum TRP (r=0.396, p=0.005), KYN/TRP ratio (r=-0.248, p=0.018), and the HAMA score (r=-0.279, p=0.008). The current data imply that decreased SpO2 during the acute phase of COVID-19 infection is predictive of anxiety caused by Long COVID. Our data reveal that around 32% of Long COVID patients have elevated IDO activity in association with elevated anxiety.

---

Acute COVID-19, particularly critical illness, is characterized by elevated activity of indoleamine-2,3-dioxygenase (IDO), the rate-limiting enzyme of the tryptophan (TRP) catabolism pathway, with decreased serum TRP and elevated kynurenine (KYN) (Almulla et al., 2022). Recent research showed a correlation between increased activity of this tryptophan catabolite (TRYCAT) pathway and anxiety symptoms caused by depression and schizophrenia (Songtachalert et al., 2018). Nevertheless, it is uncertain whether TRYCAT pathway activation contributes to anxiety caused by Long COVID. Hence, here we want to determine if TRYCAT pathway activation is linked with anxiety symptoms due to Long COVID.

All participants (n=90) had suffered from COVID-19, as shown by positive IgM and reverse transcription real-time polymerase chain reaction (rRT-PCR) to SARS-CoV-2 as well as the presence of an acute respiratory illness. In addition, we selected sixty individuals with Long COVID according to World Health Organization criteria (2021) and 30 controls without Long COVID. The 60 Long COVID patients suffered from two symptoms that impeded daily functioning for at least two months and were present 3-6 months after the acute phase of infection. Prior to participation, all patients had negative rRT-PCR tests. The Hamilton Anxiety Rating Scale (HAMA) score (Hamilton, 1959) was used to quantify anxiety severity, and a pure anxiety score including anxious mood, tension, fears, and anxiety behavior during the interview was calculated. Using ELISA kits, serum TRP and KYN levels were measured (Nanjing Pars Biochem Co., Ltd., Nanjing, China). We computed a z unit-weighted composite score as z kynurenine - z tryptophan (zKYN-zTRP). During the acute phase of infection, peripheral oxygen saturation (SpO2) was monitored using an electronic oximeter (Shenzhen Jumper Medical Equipment Co. Ltd.). SPSS version 28 (IBM, Windows version) was used for all data analysis.

Using cluster analysis and the HAMA score, the pure HAMA anxiety score, serum TRP and KYN, zKYN-zTRP (all measured during Long COVID), and SpO2 (measured during the acute phase of COVID-19), we were able to classify Long COVID patients into two distinct clusters with an adequate silhouette cohesion and separation index (0.58): cluster 1 (n=61) and cluster 2 (n=29). T**able 1** shows that cluster 2 had lower SpO2 and TRP levels, as well as higher KYN, zKYN-zTRP, and HAMA scores than cluster 1. Regression analysis revealed that the zKYN-zTRP composite explained 14.4% of the variance in the HAMA score (F=14.81, df=1/88, p=0.001). In addition, regression analysis revealed that SpO2 partially explained the variance in serum TRP (r=0.396, p=0.005), zKYN-zTRP (r=-0.248, p=0.018), and the HAMA score (r=-0.279, p=0.008). In addition, the rise in the zKYN-zTRP ratio largely mediated the effect of SpO2 on the HAMA score. The current data imply that decreased SpO2 during the acute phase of COVID-19 infection is somewhat predictive of anxiety caused by Long COVID. Lowered SpO2 is an important predictor of the severity of the immune-inflammatory response of the acute phase of COVID-19 (Al-Hadrawi et al., 2022). Our data reveal that around 32% of Long COVID patients have elevated IDO activity in association with elevated anxiety.

**Table 1:** Anxiety ratings and biomarker data of two clusters of individuals, 3-6 months after the infectious phase of COVID-19.

| **Variables** | **Cluster 1**<br>**(n=61)** | **Cluster 2**<br>**(n=29)** | **F** | **df** | **p-value** |
| --- | --- | --- | --- | --- | --- |
| Total HAMA | 11.3 (6.7) | 21.5 (10.3) | 31.28 | 1/88 | <0.001 |
| Pure HAMA | 3.6 (1.9) | 6.4 (3.4) | 25.27 | 1/88 | <0.001 |
| SpO2%* | 90.02 (2.64) | 83.72 (6.92) | 37.94 | 1/88 | <0.001 |
| L-tryptophan (ng/mL)* | 12.74 (5.69) | 9.16 (2.78) | 10.24 | 1/88 | <0.001 |
| Kynurenine (ng/mL) | 2.63 (1.20) | 5.31 (2.44) | 49.30 | 1/88 | <0.001 |
| zKYN-zTRP ratio (z score) | -0.480 (0.665) | 1.010 (0.820) | 84.79 | 1/88 | <0.001 |

Results are shown as mean ( ± SD). All results of analyses of variance (F statistic).
\*Processed in logarithmic transformation.
HAMA: Hamilton Anxiety Rating Scale, SpO2: Peripheral oxygen saturation.
zKYN-zTRP: z unit-weighted composite score computed as z L-tryptophan – z kynurenine

Decreased plasma TRP is related with greater psychological distress, depersonalization, and obsessions in major depressive disorder (Maes et al., 1990). In the early puerperium, decreased plasma TRP and increased KYN levels are associated with increased inflammatory processes and anxiety (Maes et al., 2002). Interferon-α-based immunotherapy enhances HAMA scores 2-4 weeks and 6 months following treatment initiation in hepatitis C patients in conjunction with inflammatory processes, decreased plasma TRP, higher KYN, and an enhanced KYN/TRP ratio (Bonaccorso et al., 2002).

Previously, we reviewed that the anxiogenic effects of activated immune-inflammatory pathways may be partially attributed to increased IDO activity, which results in a) decreased availability of TRP to the brain, which may affect 5-HT signaling and attenuate neuroprotection, and b) increased production of anxiogenic and neurotoxic TRYCATs, including kynurenine (Maes et al., 2011). Nevertheless, the fact that a significant proportion of the variability in TRP, zKYN-zTRP, and the intensity of anxiety remains unexplained suggests that other processes are at play. Recently, we demonstrated that Long COVID symptoms (including depression, anxiety, and fatigue) are predicted by elevated biomarkers of inflammation (C-reactive protein and the NLRP3 inflammasome), oxidative toxicity, decreased antioxidant defenses, and insulin resistance (Al-Hakeim et al., 2022a; Al-Hakeim et al., 2022b; Al-Hakeim et al., 2023).

## Data Availability

The database generated during this study will be made available from the corresponding author (MM) upon reasonable request once the data set has been fully exploited by the authors

## Acknowledgments

We thank the staff of the Dialysis Unit at Al-Hakeem general hospital and Al-Sader medical city in Najaf governorate-Iraq for their help in the collection of samples.

## Ethical approval and consent to participate

The research was approved by the University of Kufa’s institutional ethics board (8298/2022). All controls and patients as well as their guardians (parents or other close family members) gave written informed consent prior to participation in this study.

## Declaration of interest

The authors have no conflicts of interest with any industrial or other association concerning the submitted article.

## Funding

This work was supported by an FF66 grant and a Sompoch Endowment Fund from the Faculty of Medicine, MDCU (RA66/016) to MM.

## Notes

### Competing Interest Statement

The authors have declared no competing interest.

